# TRENDS IN CLINICAL STAGE AT PRESENTATION FOR FOUR COMMON ADULT CANCERS IN IBADAN, NIGERIA

**DOI:** 10.1101/2023.09.22.23295952

**Authors:** Akinyinka O. Omigbodun, Adebayo D. Agboola, Olufunke A Fayehun, Motunrayo Ajisola, Abiola Oladejo, Omolara Popoola, Richard Lilford

## Abstract

**Introduction:** Cancer outcome is largely determined by stage at diagnosis. We hypothesised that people living in Low- and Middle-Income Countries may be reaching diagnosis at an early stage, reflecting growth in awareness of the disease and its symptoms. We examined stage at diagnosis for four common cancers presenting at a referral centre in Nigeria.

**Methods:** A retrospective review of case-records from cases treated for four common cancers (colorectal, uterine cervix, breast and prostate) at University College Hospital, Ibadan over two epochs: 2012-2013 and 2017-2018.

**Results:** There is no evidence that cases were diagnosed at an earlier stage in the later epoch. Seventy-five percent of cases were diagnosed at late stage (III or IV) in the second epoch, vs seventy percent in the first epoch.

**Conclusion:** There is no evidence in the data that the problem of late presentation of common cancers is reducing in Oyo State, Nigeria.

## INTRODUCTION

Cancer is a global epidemic and one of the leading causes of death of one in six mortality worldwide.(1-4) More than half of new cancer cases and two-thirds of cancer deaths occur in low-and-middle-income countries (LMICs).(5-7)

Reducing delay in between first symptom and treatment is a high priority for LMICs where cases are often at an advanced stage by the time they are diagnosed.(8-15) There is extensive evidence that earlier diagnosis can considerably reduce the mortality and costs of cancer and improve quality of life.(16-18) It might be hypothesised that cancers are diagnosed at increasingly early stages in LMICs as people move to cities and as the population becomes better educated. The purpose of this paper is to test this hypothesis with respect to one LMIC location in Nigeria.

We describe stage at presentation for four common adult cancers presenting at a tertiary hospital in 2012-2013 and 2017-2018 in Ibadan, South West Nigeria.

## METHODS

### Study Design

This descriptive study was based on retrospective data of histologically diagnosed breast, cervical, prostate and colorectal cancer patients at the University College Hospital (UCH) in Ibadan, with an analytical sample of 631 patients over two, biennial, epochs (2012-2013 and 2017-2018). We analysed data from the Cancer Registry at UCH about patients from the Surgery, Gynaecology and Radiotherapy departments. The hospital is a referral centre for patients needing cancer care within and outside Ibadan.

### Data Collection

We retrieved hospital case files with follow-up records and radiotherapy treatment cards from January 2012 to December 2013 and January 2017 to December 2018 for patients with breast, cervical, prostate, and colorectal cancer. We obtained information on age, parity, educational status, employment status, comorbidity, stage of cancer, treatment received, response from treatment and follow-up status from the case files.

### Variable measurements

Breast,(19) prostate,(20) and colorectal (21) cancers were staged using the American Joint Committee on Cancer staging, while cervical (22) cancer was staged using the International Federation of Gynaecology and Obstetrics (FIGO) staging system. The stages were grouped into I, II, III and IV. The study explores the association of education, distance to health facility and mode of referral to stage at presentation over the above two epochs. The highest educational level was categorised into primary, secondary and tertiary. The distance grouping to the health facility was based on approximately 50 kilometres to the tertiary health facility.

### Data analysis

For this descriptive study, we examined the proportion of diagnoses that were late (stages III or IV) over the two epochs with respect to all cancers and the individual cancers. For reasons that will become apparent, we made the comparison for ‘all’ cancers with and without including cervical cancers. We also examined education and distance to the health facility by stage at diagnosis and type of cancer presented over the two study epochs. The results were presented in the form of frequency distribution and graphs. The data was analysed using SPSS version 20.0.

### Ethical considerations

Ethical approval was obtained from the University of Ibadan / University College Hospital Ethical Review Committee with number UI/EC/19/0633. All the information and data retrieved from the patient case files were treated with utmost confidentiality, and identifiers that could be traced to participants were removed.

## RESULTS

### Numbers and types of cancer

There were 248 and 383 histologically confirmed cervical, breast, prostate, and colorectal cancer cases at the tertiary hospital in 2012-2013 and 2017-2018, respectively. Table 1 shows the numbers and proportions of cancer types over the two study epochs. Breast cancer was the most common type of cancer over both epochs. The prevalence of cervical cancer increased by a greater proportion than the other cancers.

**Table 1:** Distribution of cancer types by epoch.

| Types of cancer | 2012-2013 |  | 2017-2018 |  |
| --- | --- | --- | --- | --- |
|  | % | n | % | n |
| Breast | 44.8 | 111 | 33.2 | 127 |
| Cervical | 24.6 | 61 | 34.2 | 131 |
| Prostate | 19.7 | 49 | 21.9 | 84 |
| Colorectal | 10.9 | 27 | 10.7 | 41 |
| Total | 100.0 | 248 | 100.0 | 383 |

In both epochs, prostate and colorectal cancers accounted for at least 19% and 10% of the diagnoses, respectively.

### Stage at presentation

Nearly 70% (69.8%) of patients with our index cancers presented at late stage (III or IV) in epoch 1, while 74.6% presented late in epoch 2 (Figure 1a). There is no evidence of a shift to a lower proportion of late-stage diagnoses in the second epoch compared to the first, and the trend was in the opposite direction (Figure 1a Mann-Whitney z=-1.383 [p=0.167]). Figure 1b shows the same data excluding cervical cancer, for reasons given in the Discussion. Again, there is no visual or statistical evidence of a trend to earlier diagnosis in the later epoch (Mann-Whitney z=-1.082 [p=0.279]).

**Figure 1a:**
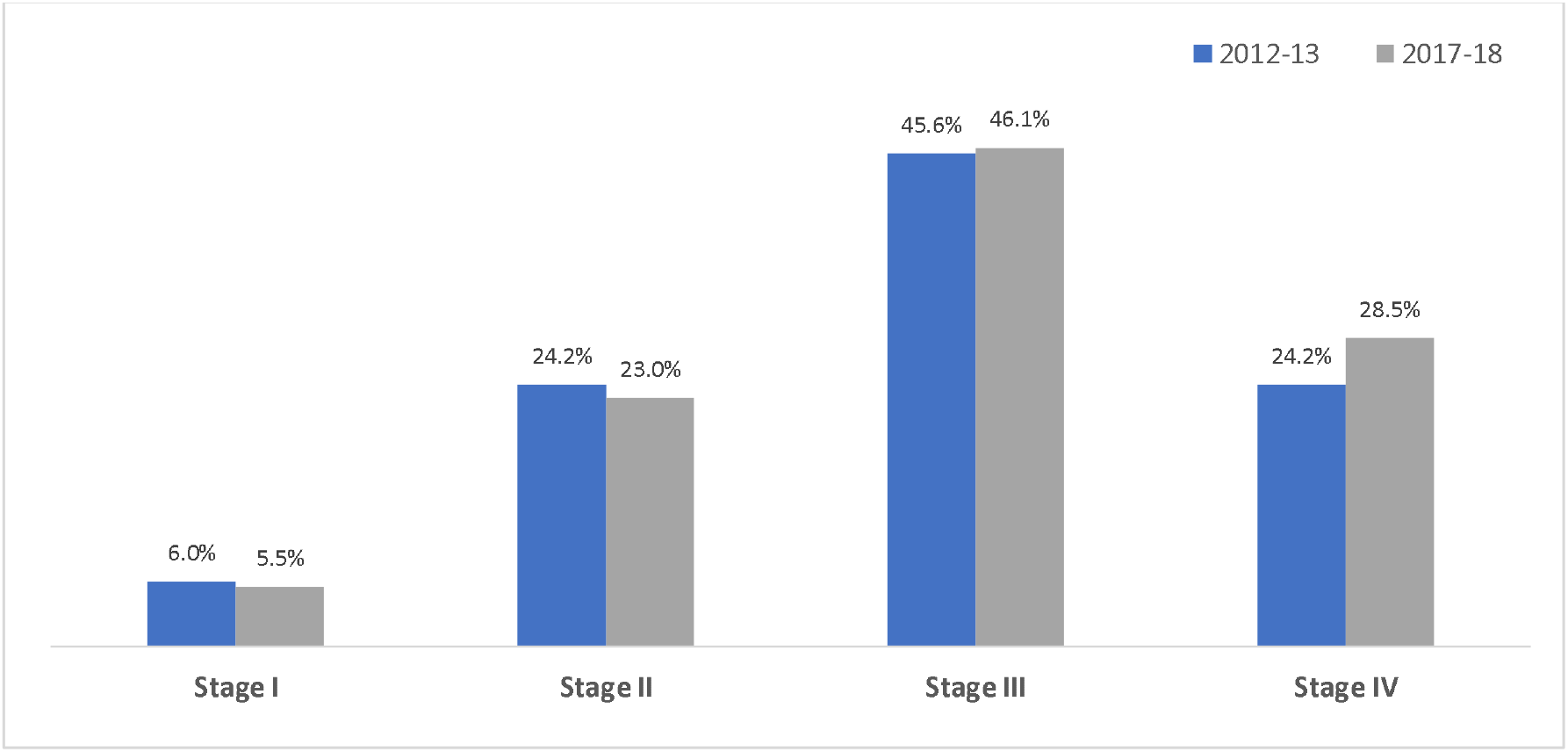
Stage at presentation of four cancer types

**Figure 1b:**
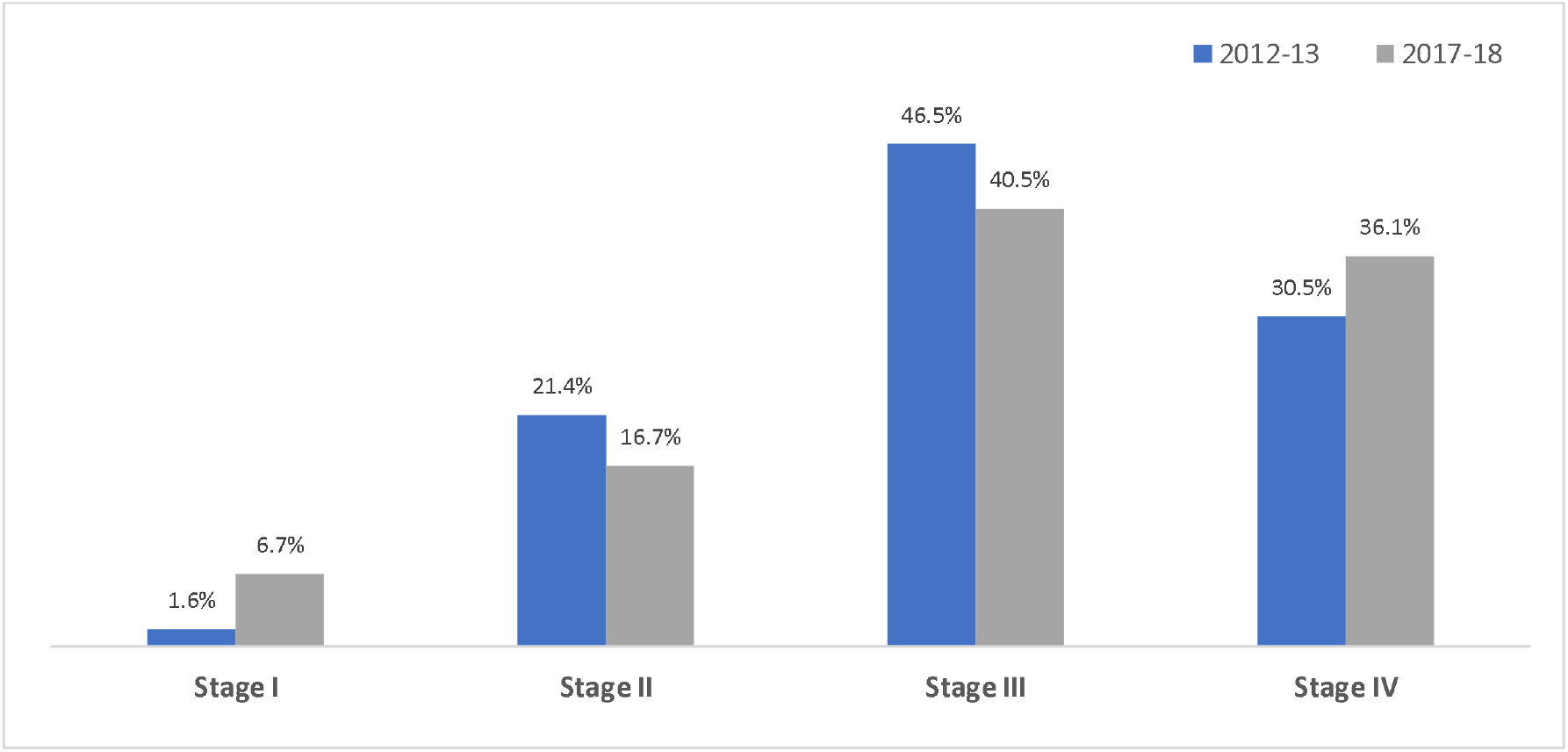
Stage at presentation of three cancer types (excluding cervical cancer)

With regard to individual cancer types, 87.4% of breast cancers presented late in the early epoch, decreasing to 80.1% in the later epoch. However, the trend was reversed for prostate cancer (increasing from 61.3% to 72.6%), for colorectal cancer (increasing from 60.3% to 70.7% and for cervical cancer (increasing from47.5% to 61.8%). More detailed breakdown by stage is given in Table 2.

**Table 2:** Association between stage at presentation and cancer type.

| Epoch | Type of cancer | Cancer stage at presentation (%) |  |  |  | Chi-square; p-value* |
| --- | --- | --- | --- | --- | --- | --- |
|  |  | Stage I | Stage II | Stage III | Stage IV |  |
| 2012-2013 | Breast | 0.0 | 12.6 | 55.9 | 31.5 | 56.26; 0.000 |
|  | Cervix | 19.7 | 32.8 | 42.6 | 4.9 |  |
|  | Prostate | 2.0 | 36.7 | 32.7 | 28.6 |  |
|  | Colorectal | 7.4 | 29.6 | 33.3 | 29.6 |  |
| 2017-2018 | Breast | 4.7 | 14.2 | 51.2 | 29.9 | 45.20; 0.000 |
|  | Cervix | 3.1 | 35.1 | 48.1 | 13.7 |  |
|  | Prostate | 8.3 | 19.0 | 27.4 | 45.2 |  |
|  | Colorectal | 9.8 | 19.5 | 34.1 | 36.6 |  |

*\* Statistical analysis within epoch*

### Factors affecting stage of presentation across both epochs

An examination of education and distance to health facility for all cancer types is presented in Table 3.

**Table 3:** Education, distance to health facility and mode of referral on time to cancer presentation in a tertiary hospital.

| Epoch | Characteristics | Cancer stage at presentation (%) |  |  |  | Chi-square;<br>p-value* |
| --- | --- | --- | --- | --- | --- | --- |
|  |  | Stage I | Stage II | Stage III | Stage IV |  |
| 2012-2013 | *Education* |  |  |  |  | 9.05; 0.171 |
|  | Primary (*n*=69) | 4.3 | 15.9 | 53.6 | 26.1 |  |
|  | Secondary (*n*=98) | 8.2 | 23.5 | 48.0 | 20.4 |  |
|  | Tertiary (*n*=81) | 4.9 | 32.1 | 35.8 | 27.2 |  |
|  | *Distance to HF* |  |  |  |  | 2.65; 0.449 |
|  | Within 50km (*n*=227) | 6.2 | 22.9 | 46.7 | 24.2 |  |
|  | Outside 50km (*n*=21) | 4.8 | 38.1 | 33.3 | 23.8 |  |
| 2017-2018 | *Education* |  |  |  |  | 11.86; 0.065 |
|  | Primary (*n*=85) | 5.9 | 17.6 | 44.7 | 31.8 |  |
|  | Secondary (*n*=177) | 3.4 | 26.6 | 47.5 | 22.6 |  |
|  | Tertiary (*n*=121) | 8.3 | 21.5 | 35.5 | 34.7 |  |
|  | *Distance to HF* |  |  |  |  | 6.68; 0.083 |
|  | Within 50km (*n*=270) | 5.2 | 19.6 | 44.4 | 30.7 |  |
|  | Outside 50km (*n*=113) | 6.2 | 31.0 | 39.8 | 23.0 |  |

\* Statistical analysis within epoch

Compared with patients with more advanced education, patients with low levels of education presented at a more advanced stage in the tertiary health facility for both epochs. Distance to the health facility was not significantly associated with the stage of presentation; about 70% of patients living within 50km of the hospital were histologically diagnosed at stages III and IV.

The time to presentation of cancer in tertiary hospital was further examined by removing patients with cervical cancer for reasons mentioned in the Discussion. The pattern of late presentation at the tertiary hospital, as shown in Table 4 for the breast, prostate and colorectal, did not differ from when cervical cancer was included in the dataset.

**Table 4:**
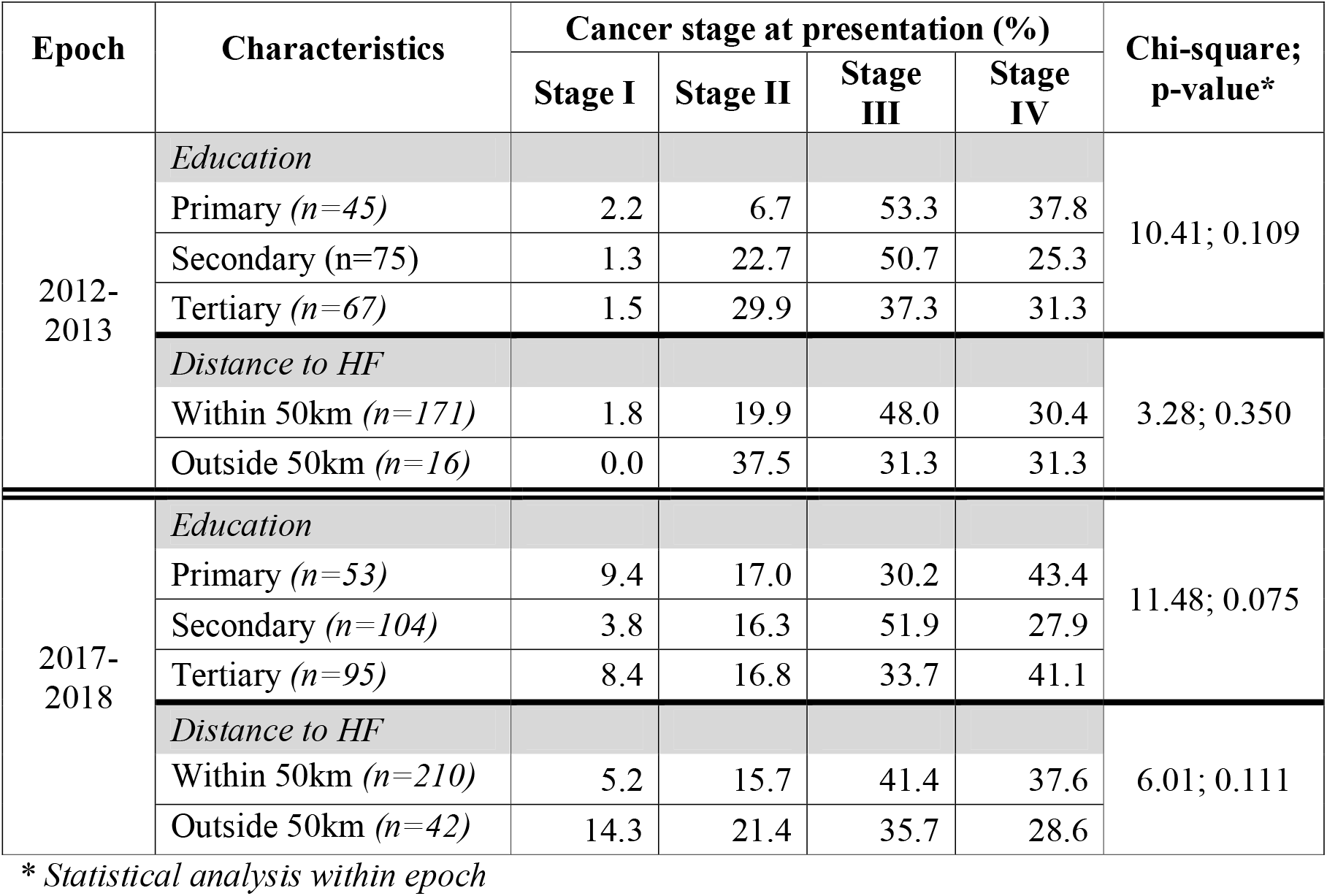
Education, distance to health facility and mode of referral on time to breast, prostate and colorectal cancers presentation in a tertiary hospital.

| Epoch | Characteristics | Cancer stage at presentation (%) |  |  |  | Chi-square;<br>p-value* |
| --- | --- | --- | --- | --- | --- | --- |
|  |  | Stage I | Stage II | Stage III | Stage IV |  |
| 2012-2013 | *Education* |  |  |  |  | 10.41; 0.109 |
|  | Primary (n=45) | 2.2 | 6.7 | 53.3 | 37.8 |  |
|  | Secondary (n=75) | 1.3 | 22.7 | 50.7 | 25.3 |  |
|  | Tertiary (n=67) | 1.5 | 29.9 | 37.3 | 31.3 |  |
|  | *Distance to HF* |  |  |  |  | 3.28; 0.350 |
|  | Within 50km (n=171) | 1.8 | 19.9 | 48.0 | 30.4 |  |
|  | Outside 50km (n=16) | 0.0 | 37.5 | 31.3 | 31.3 |  |
| 2017-2018 | *Education* |  |  |  |  | 11.48; 0.075 |
|  | Primary (n=53) | 9.4 | 17.0 | 30.2 | 43.4 |  |
|  | Secondary (n=104) | 3.8 | 16.3 | 51.9 | 27.9 |  |
|  | Tertiary (n=95) | 8.4 | 16.8 | 33.7 | 41.1 |  |
|  | *Distance to HF* |  |  |  |  | 6.01; 0.111 |
|  | Within 50km (n=210) | 5.2 | 15.7 | 41.4 | 37.6 |  |
|  | Outside 50km (n=42) | 14.3 | 21.4 | 35.7 | 28.6 |  |

\* Statistical analysis within epoch

## DISCUSSION

The prevalence of all four cancer types increased between the two epochs. Not only did cancers tend to present in late stage, but the proportions presenting late did not decline over the epochs – if anything, the trend was towards an increase. Only for breast cancer was there a small reduction (six percentage points) in cancer stage. There is thus no signal that patients are presenting earlier, or being more rapidly diagnosed when they present to health services. It is unlikely that much improvement in outcome will be seen at a population level while patients continue to present late. Therapy to treat advanced cancer is improving, but it is expensive and not widely available to the majority of patients in LMICs and the largest gains can be achieved by early diagnosis and treatment.

Our study does have limitations. Most crucially, it is not a population-based study and is therefore influenced by changes in population structure, and also by changing rates of referral. Indeed, the University College Hospital, Ibadan installed new radiotherapy equipment between the two epochs, which attracted cervical cancer referrals from distant hospitals. This is reflected in the patient numbers with cervical cancer showing the greatest increase. It was for this reason that we analysed data with and without including cervical cancer. It is nevertheless possible that, with respect to cervical cancer, a tendency to earlier diagnosis was disguised by referral of later presentation cases from other hospitals. If so, this finding is not confirmed by the stage at presentation for the other three cancer types.

Our study was conducted in only one hospital in one country. There is nothing about this setting to suggest that it is unique with respect to cancer diagnosis. That said, a population-based study over multiple countries would be desirable. The possibility of such a study is limited by the incomplete development of reliable cancer registries in many LMICs.

Our findings reinforce the need for more research into interventions to reduce waiting times and thereby ‘down-stage’ cancers at the time of diagnosis. An expert group recently voted research into reducing cancer delay the top priority for research across all of cancer care.(23)

## CONCLUSION

There is an increasing trend in the prevalence of cancers globally, and this study revealed that Nigeria is not exempt. The proportion of patients presenting with late stages of four common adult cancers in Southwest Nigeria is not declining.

## Data Availability

All data produced in the present work are contained in the manuscript. Data used are held by the University College Hospital, Ibadan, Nigeria.

## ACKNOWLEDGEMENTS

We thank Peter Chilton, University of Birmingham, for their help in editing and formatting the manuscript.

